# Not Too Late to Intervene? A Meta-analysis of 13 Studies Evaluating the Association of Endovascular Therapy with Clinical Outcomes in Stroke Patients Presenting Beyond 24 Hours

**DOI:** 10.1101/2024.09.03.24313005

**Authors:** Mohamed F Doheim, Abdulrahman Ibrahim Hagrass

## Abstract

**Background:** Association of endovascular therapy (EVT) with clinical outcomes beyond 24 hours remains unclear. We conducted a meta-analysis to answer this question.

**Methods:** We searched for eligible studies in PubMed from inception until June 2023. The outcomes included functional independence, as assessed with 90-day modified Rankin Scale (mRS) scores (0-2), thrombolysis in cerebral infarction (TICI) scores (2b-3 or 3), symptomatic intracranial hemorrhage (sICH), and 90-day mortality. Risk ratio (RR) and 95% confidence interval (CI) were pooled.

**Results:** We finally included 13 studies in our meta-analysis (number of patients treated with EVT beyond 24 h=866). For single arm analysis, the pooled estimates of functional independence (mRS 0-2), sICH, and mortality were 0.342 (95% CI = 0.275 -0.410, P < 0.001), 0.062 (95% CI = 0.045 -0.078, P < 0.001), and 0.232 (95% CI = 0.164 -0.301, P < 0.001); respectively with successful reperfusion (TICI 2b-3) of 0.837 (95% CI = 0.812 -0.861, P < 0.001). Comparing EVT with medical management, the pooled analysis showed that EVT had a statistically significant advantage over medical management (RR = 2.62, 95% CI [1.38, 4.96], P = 0.003). However, our analysis showed a higher incidence of sICH in EVT group (RR = 3.58, 95% CI [1.53, 8.37], P = 0.003). When we pooled studies comparing EVT beyond 24 h with EVT within 6–24 h, the findings showed no statistically significant difference for functional independence, sICH, and 90-d Mortality.

**Conclusion:** EVT is associated with better clinical outcomes than medical management beyond 24 hours. These results are iconoclastic enhancing a new paradigm in which a contemporary restriction to specific time window to treat patients rather than their own clinical and imaging characteristics seems to be anecdotal. Prospective studies are needed to confirm the best eligible patients for EVT in this newly proposed window extension.

## Introduction

It was not until 2015 that endovascular therapy (EVT) was established as a standard treatment for anterior circulation large vessel occlusions (LVOs) in the early window following the publications of five pivotal randomized controlled trials (RCTs) that were subsequently pooled in a patient level data-meta-analysis.^1^ In 2018, two imperative RCTs (DAWN (Thrombectomy 6 to 24 Hours After Stroke With a Mismatch Between Deficit and Infarct trial) and DEFUSE 3 (Thrombectomy for Stroke at 6 to 16 Hours With Selection by Perfusion Imaging trial)) demonstrated that the EVT can be performed within 6-24 hours in selected patients.^2, 3^ In AURORA patient level data meta-analysis, the pooled results of six published RCTs confirmed these results.^4^

Recently, a multicenter cohort study assessed extended window patients not fulfilling the DEFUSE-3 and DAWN inclusion criteria and found EVT to be more associated with favorable functional outcomes compared to medical management.^5^ Additionally, recent studies questioned the possible role of EVT beyond 24 hours with promising results.^6-8^ In this systematic review and meta-analysis, we aimed to evaluate the current evidence and provide pooled estimate on study level data reporting EVT beyond 24 hours.

### Methods Study Design

This review was conducted following the recommendations provided by the Preferred Reporting Items for Systematic review and Meta-Analysis (PRISMA) ^9^.

### Eligibility Criteria and Study Selection

We included clinical studies that reported functional and safety outcomes in patients who were treated with EVT beyond 24 hours following ischemic strokes. Our criteria for excluding studies were review articles, non-English studies, and conference abstracts.

### Database and Search Strategy

We used PubMed to search the Midline database thoroughly for articles up until February 2023. For our search, we utilized this search strategy: (endovascular OR vascular surgical OR thrombectomy OR thromboaspirat* OR “thrombectomy”) AND (stroke OR brain ischemia OR cerebrovascular OR brain vascular OR “stroke”[MeSH Terms]) AND (Very Late OR late OR 24-hr OR 24 hr OR 24 Hours OR 24-Hours).

### Quality Assessment

To evaluate the quality of the included cohort studies, we used the NIH Quality Assessment Tool for Observational cohort/cross-sectional studies ^10^. While the NIH Quality Assessment Tool for case series/case reports was used to evaluate the case series studies ^11^.

## Data Extraction

Data from the chosen studies was extracted and placed into a Microsoft Excel spreadsheet covering the following topics: 1) Summary of included studies: author, publication year, design, time period, and inclusion criteria; 2) Baseline characteristics of the included population: age, gender, hypertension, diabetes mellitus, admission NIHSS score and occlusion location; and 3) Study outcomes: primary outcome (functional independence at 90 days); and secondary outcomes (symptomatic intracranial hemorrhage (sICH), 90-day mortality, Thrombolysis in Cerebral Infarction (TICI) 2b-3, and TICI 3).

## Data Synthesis

Throughout the process of analyzing the data, we made use of two different software programs. We used “Open Meta-Analyst” to do a meta-analysis with a single treatment group. While we ran the meta-analysis of the two-arm studies with Review Manager version 5.4 (The Cochrane Collaboration, Oxford, United Kingdom). The risk ratio (RR) and 95% confidence interval (CI) were calculated using dichotomous data from the included studies. A p-value of less than 0.05 would have been considered statistically significant. The degree of heterogeneity across groups was measured with the χ2 and I-square (I2). Since the χ2 P value was less than 0.1 or the I2 was >50%, we concluded that the data was not homogeneous. We used the random-effect model when there was evidence of heterogeneity and the fixed-effect model otherwise.

## Results Literature search

Figure 1 displays the results of the search and selection process employed in our literature review. The original search turned up 4,679 entries, and after discarding 1816 duplicates, we were left with 2,863 records. We started by reviewing the abstracts and titles of all 2863 papers, and then proceeded on to the complete texts of 54 of those publications. The final analysis consisted of a total of 13 papers ^6, 12-23^, all of which were included because they satisfied our inclusion criteria.

**Figure 1:**
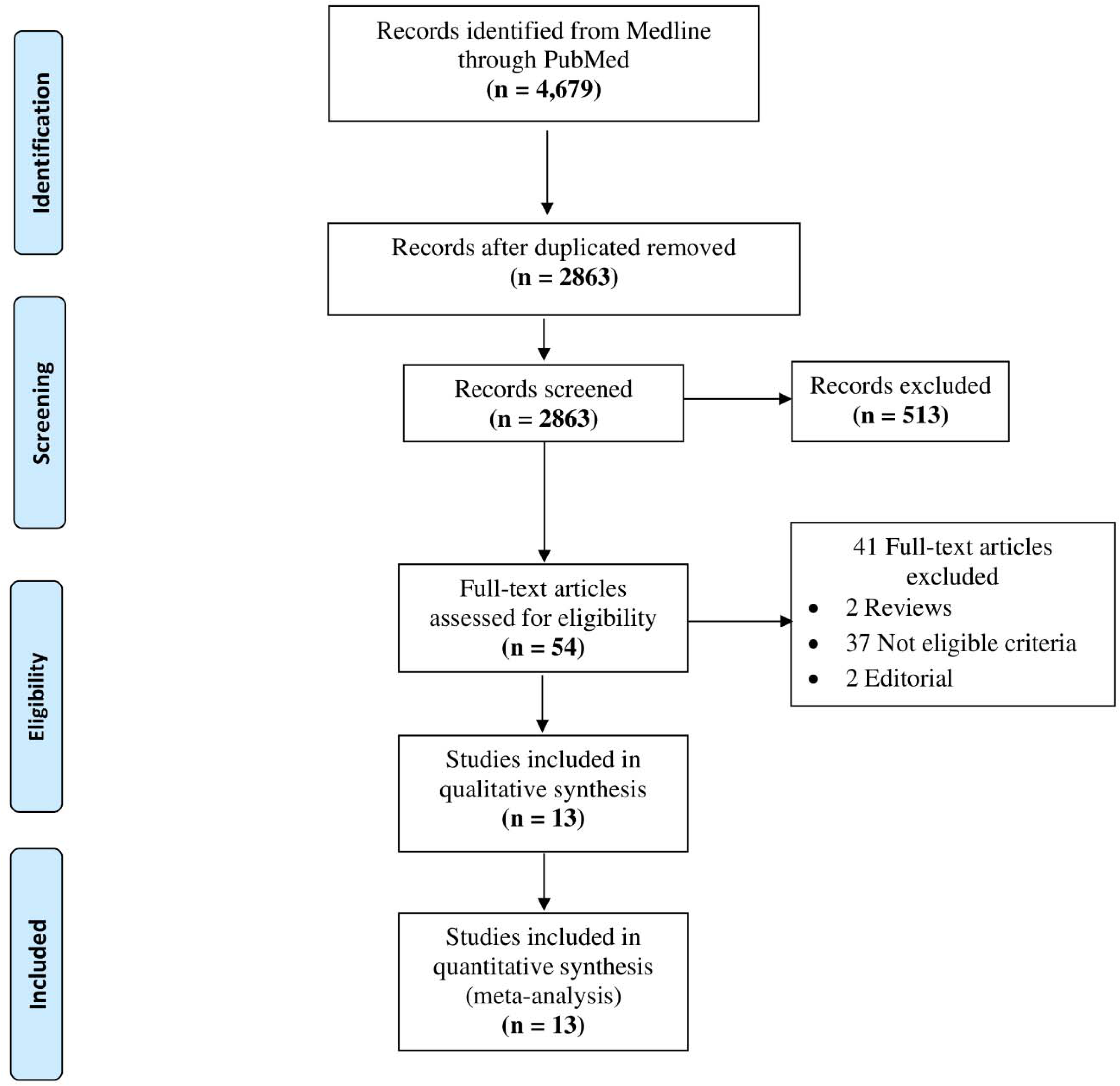
PRISMA flow diagram.

### Characteristics of the included studies & populations’ baseline

The number of participants who underwent thrombectomy after 24 hours, thrombectomy within 6–24 hours, and those who received medical management were 866, 5543, and 262 respectively. The age distributions of those who underwent thrombectomy after 24 hours, thrombectomy within 6–24 hours, and those who received medical management ranged from 65 to 75, 57 to 69.17, and 64 to 78.3 years, respectively. There were 458 males (52.9% of the total) in the group that underwent thrombectomy more than 24 hours after the onset of symptoms, 2756 males (49.7% of the total) in the group that underwent thrombectomy between 6 and 24 hours, and 119 males (45.4%) in the medical management group. The control arm reported by Purrucker et al. included all patients who received treatment within 24 hours, without providing distinct data for those treated within the 6–24-hour timeframe ^19^. The most relevant aspects of the studies used in this meta-analysis are summarized in Tables 1 and 2.

**Table 1:** Summary of included studies.

| Study ID | Type | Time period | EVT after 24 h | Number EVT within 6–24 h | Medical Management | Main selection criteria |
| --- | --- | --- | --- | --- | --- | --- |
| Desai 2018 | Retrospective of 3 centers | 2010 – 2018 | 21 | N/A | N/A | >24 h after LSW<br>pmRS 0–1<br>Age < 80:<br>NIHSS ≥ 10 + core < 31 ml<br>or NIHSS ≥ 20 + core < 51 ml;<br>age ≥ 80:<br>NIHSS ≥ 10 + core < 21 ml |
| Purrucker 2022 | Retrospective analysis of prospectively acquired data, single center | 2014 – 2021 | 43 | 2304 | N/A | >24 h after LSW or >24 h after definite onset of symptoms; Patients treated <24h served as a group for comparison |
| Dhillon 2022 | Prospective cohort | 2015 – 2020 | 104 | 1046 | N/A | >24 h after LSW or onset of symptoms; Patients treated within 6-24 hours served as a control group |
| Shaban 2022 | Retrospective analysis of prospectively maintained multicenter registry Stroke Thrombectomy and Aneurysm Registry. | 2013 – 2021 | 121 | 1821 | N/A | >24 h after LSW; Patients treated within 6-24 hours served as a control group |
| Sarraj 2022 | Retrospective cohort | 2012 – 2021 | 185 | N/A | 116 | >24 h after LSW |
| Casetta 2021 | Retrospective analysis of prospective national registry | NR | 34 | N/A | N/A | >24 h after onset of symptoms<br>NIHSS ≥ 6, collateral-score 2–3,<br>CTP core (CBV) ≥ 50% (MTT) or <1/3 of MCA territory; pmRS 0–2 |
| Manning 2018 | Retrospective analysis of prospective registry at 2 CSCs | 2016 – 2017 | 5 | N/A | N/A | >24 h after onset of symptoms |
| Mokin 2018 | Retrospective registry analysis | NR | 3 | 268 | N/A | Beyond 6 hours of stroke symptom onset (either within 6-24h or >24h) |
| Iezzi 2023 | Prospective cohort | 2019 – 2022 | 17 | N/A | 10 | >24 h after LSW |
| Pandhi 2023 | Retrospective analysis of prospective registry | 2018 – 2022 | 39 | N/A | N/A | >24 h after LSW or onset of symptoms |
| Ha 2023 | Retrospective analysis of prospective registry | 2016 – 2019 | 61 | 104 | N/A | Patients who underwent EVT in an emergency setting |
| Mohamed 2023 | Retrospective analysis of 11 stroke centers | 2015 - 2021 | 214 | N/A | 120 | >24 h after LSW |
| Dhillon 2023 | Retrospective analysis of a single center | 2018-2022 | 19 | N/A | 16 | >24 h after LSW<br>Alberta Stroke Program Early CT Score of ≥5<br>National Institutes of Health Stroke Scale of ≥6 |

CSC: comprehensive stroke center, EVT: endovascular therapy, LSW: last seen well, N/A: not applicable, NR: not reported

**Table 2:**
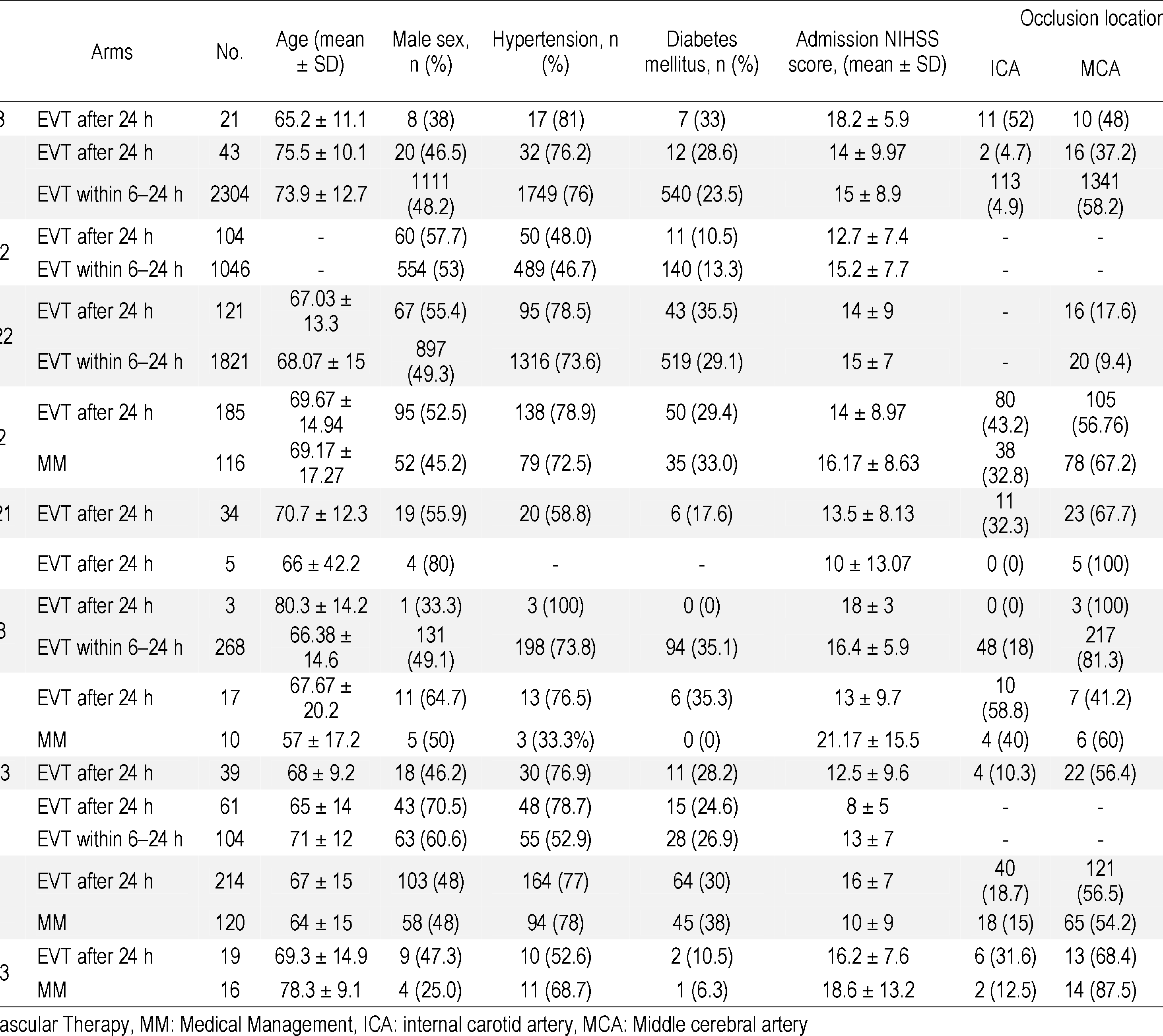
seline for included studies.

### Quality assessment

Using the NIH Quality Assessment Tool for Observational Cohort/Cross-Sectional Studies, all but Dhillon et al. and Mohamed et al. which obtained a fair grade, were of good quality ^18, 22^. On the other hand, the included case series studies were all of good quality. (Tables 3 & 4)

### Single-armed analysis

#### Functional independence at 90 days (mRs 0-2)

Our analysis of functional independence included 693 patients, who were obtained from 12 studies ^6, 12-17, 19-23^. The pooled estimate was 0.342 (95% CI = 0.275 -0.410, P < 0.001) with considerable heterogeneity (P < 0.001, I2 = 65.5%). (Figure 2a)

**Figure 2:**
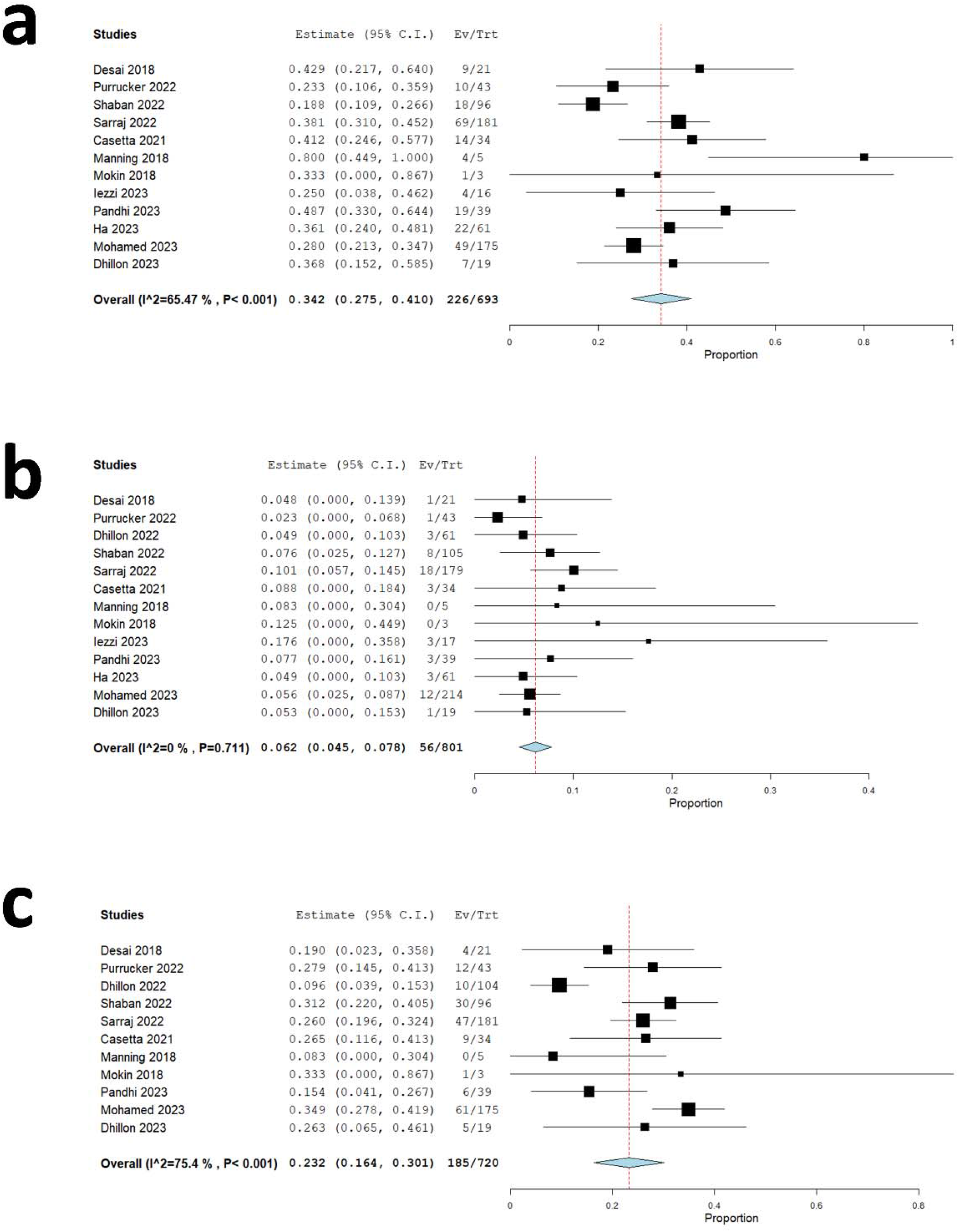
Forest plot of single-arm meta-analysis for a) Functional independence at 90 days; b) Symptomatic ICH; c) 90-d Mortality.

#### sICH

Our analysis of sICH included 801 patients, who were obtained from 13 studies ^6, 12-23^. The pooled estimate was 0.062 (95% CI = 0.045 -0.078, P < 0.001) with no evidence of heterogeneity in the combined data (P = 0.711, I2 = 0%). (Figure 2b)

#### 90-d Mortality

Our analysis of 90-d mortality included 720 patients, who were obtained from 11 studies ^6, 12-19, 22, 23^. The pooled estimate was 0.232 (95% CI = 0.164 -0.301, P < 0.001) with considerable heterogeneity (P < 0.001, I2 = 75.4%). (Figure 2c)

#### TICI 2b-3

Our analysis of TICI 2b-3 included 850 patients, who were obtained from 13 studies ^6, 12-23^. The pooled estimate was 0.837 (95% CI = 0.812 -0.861, P < 0.001) with no evidence of heterogeneity in the combined data (P = 0.209, I2 = 0%). (Figure 3a)

**Figure 3:**
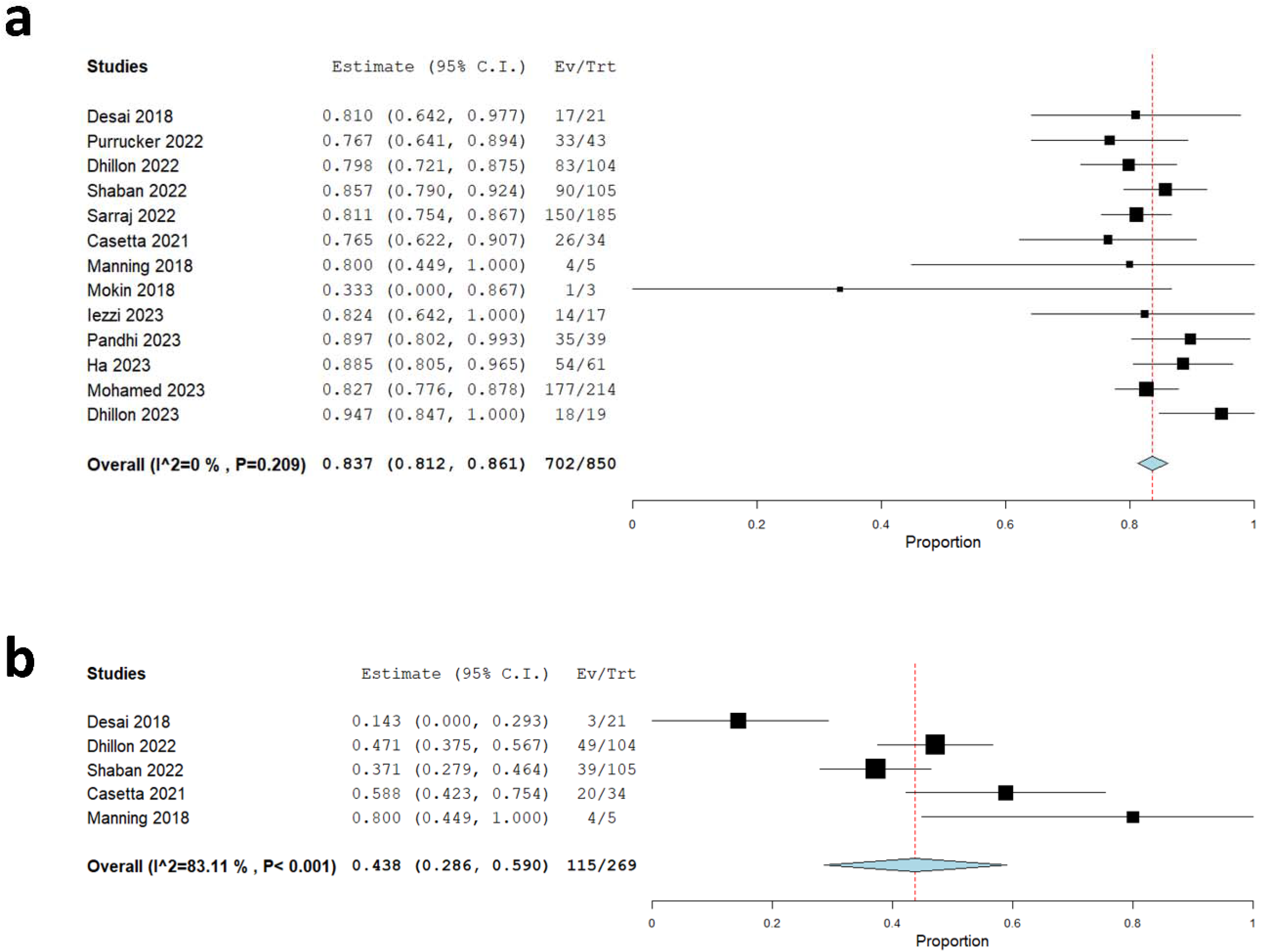
Forest plot of single-arm meta-analysis for a) TICI 2b-3; b) TICI 3.

#### TICI 3

Our analysis of TICI 3 included 269 patients, who were obtained from five studies ^6, 12, 13, 17, 18^.

The pooled estimate was 0.438 (95% CI = 0.286 -0.590, P < 0.001) with considerable heterogeneity (P < 0.001, I2 = 83.1%). (Figure 3b)

### Thrombectomy vs Medical Management

#### Functional independence at 90 days (mRs 0-2)

In total, our meta-analysis of functional independence included 616 patients, who were obtained from four studies ^16, 20, 22, 23^. The findings of the analysis showed a non-statistically significant difference between late thrombectomy and medical management (RR = 1.55, 95% CI [0.52, 4.56], P = 0.43) with marked heterogeneity in the combined data (P < 0.001, I2 = 89%) (Figure 4a). The heterogeneity can be resolved by excluding Mohamed et al.^22^, and the results showed that late thrombectomy had a statistically significant advantage over medical management (RR = 2.62, 95% CI [1.38, 4.96], P = 0.003), with no evidence of heterogeneity in the combined data (P = 0.26, I2 = 25%).

**Figure 4:**
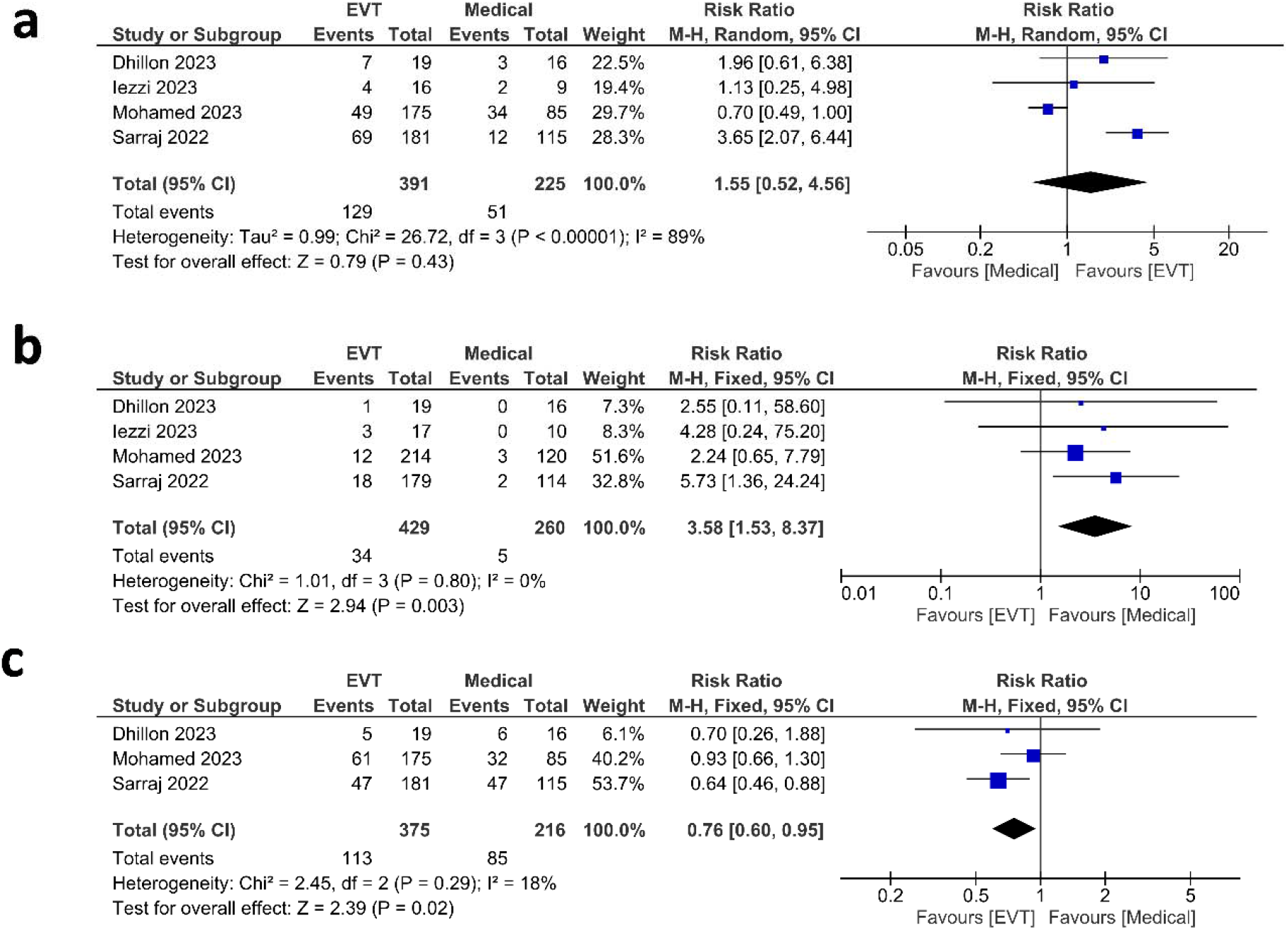
Forest plot of meta-analysis in Thrombectomy vs Medical Management groups for a) Functional independence at 90 days; b) Symptomatic ICH c) 90-d Mortality.

#### sICH

Our analysis of sICH included 689 patients, who were obtained from four studies ^16, 20, 22, 23^. Our analysis showed a higher incidence of symptomatic ICH in late thrombectomy group (RR = 3.58, 95% CI [1.53, 8.37], P = 0.003) with no evidence of heterogeneity in the combined data (P = 0.80, I2 = 0%). (Figure 4b)

#### Mortality at 90 days

Our analysis of 90 days mortality included 591 patients, who were obtained from three studies ^16, 22, 23^. The findings of the analysis showed a lower mortality rate in late thrombectomy group (RR = 0.76, 95% CI [0.60, 0.95]), P = 0.02) with no heterogeneity in the combined data (P = 0.29, I2 = 18%). (Figure 4c)

### Thrombectomy beyond 24 h vs thrombectomy within 6–24 h

#### Functional independence at 90 days (mRs 0-2)

Our analysis of functional independence included 2827 patients, who were obtained from four studies ^15, 17, 19, 21^. The finding of the analysis showed no significant difference (RR = 0.77, 95% CI [0.44, 1.36], P = 0.37) with considerable heterogeneity (P = 0.02, I2 = 71%) (Figure 5a). After excluding Ha et al. ^21^, the results showed that thrombectomy within 6-24 h had a statistically significant advantage over thrombectomy beyond 24 h (RR = 0.57, 95% CI [0.40, 0.80], P = 0.001) with no evidence of heterogeneity in the combined data (P = 0.92, I2 = 0%). (Figure 5b)

**Figure 5:**
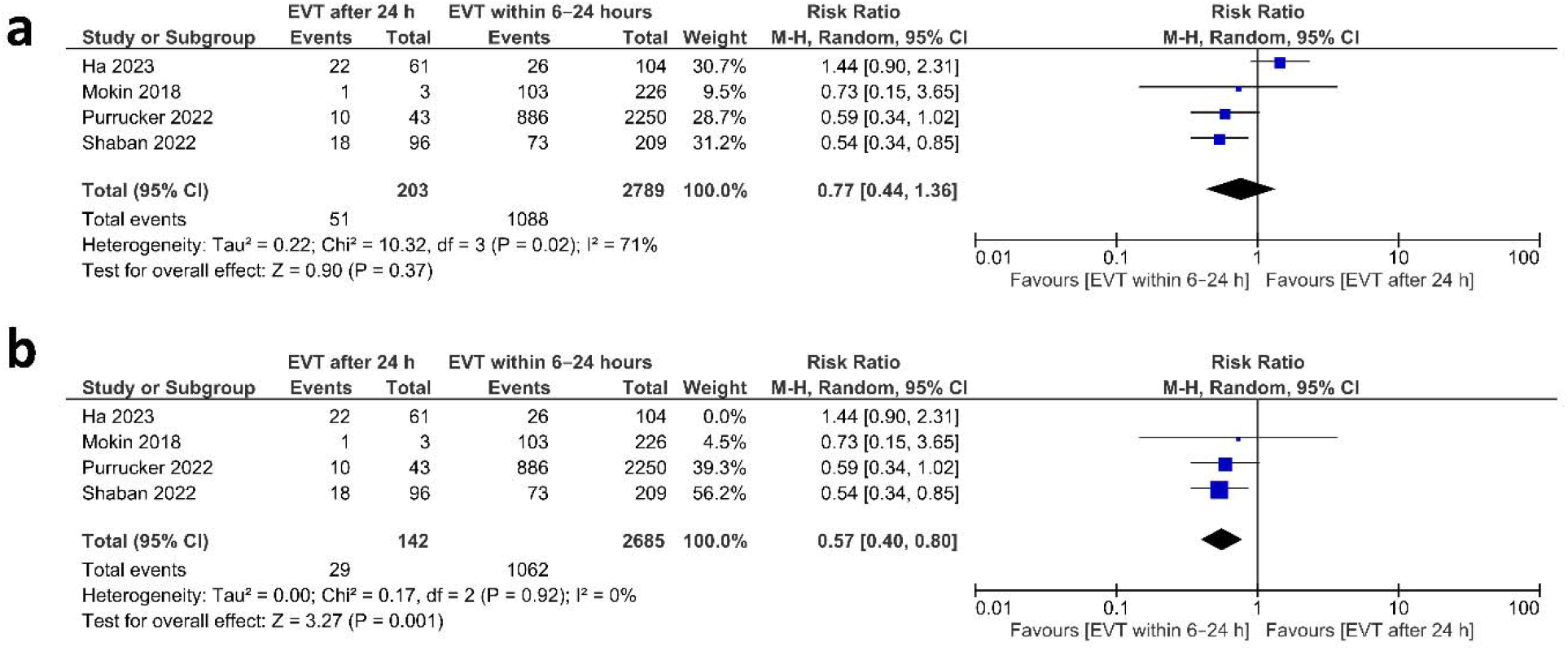
Forest plot of meta-analysis in Thrombectomy beyond 24 h vs thrombectomy within 6–24 h for a) Functional independence at 90 days; b) Functional independence at 90 days after sensitivity analysis.

#### sICH

Our analysis of symptomatic ICH included 3269 patients, who were obtained from five studies ^15, 17-19, 21^. The findings of the analysis showed no statistically significant differences between thrombectomy within 6-24 h and thrombectomy beyond 24 h (RR = 0.84, 95% CI [0.47, 1.48], P = 0.54) with no evidence of heterogeneity in the combined data (P = 0.56, I2 = 0%). (Figure 6a)

**Figure 6:**
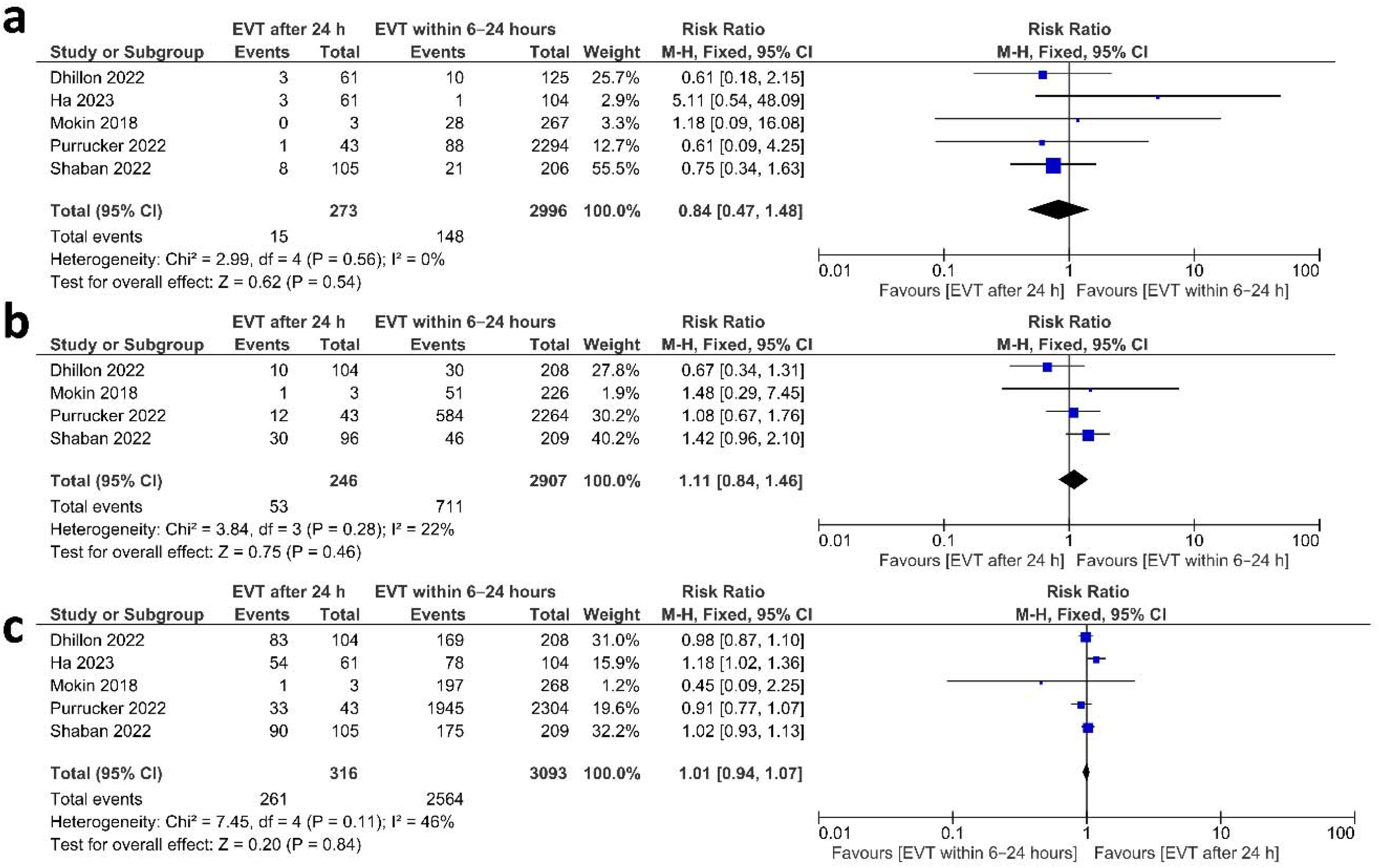
Forest plot of meta-analysis in Thrombectomy beyond 24 h vs thrombectomy within 6–24 h for a) Symptomatic ICH b) 90-d Mortality; c) TICI 2b-3.

#### 90-d Mortality

Our analysis of 90-d Mortality included 3153 patients, who were obtained from four studies ^15, 17-19^. The findings of the analysis showed no statistically significant differences between thrombectomy within 6-24 h and thrombectomy beyond 24 h (RR = 1.11, 95% CI [0.84, 1.46], P = 0.46) with mild heterogeneity in the combined data (P = 0.28, I2 = 22%). (Figure 6b)

#### TICI 2b-3

Our analysis of TICI 2b-3 included 3409 patients, who were obtained from five studies ^15, 17-19, 21^. The findings of the analysis showed no statistically significant differences between thrombectomy within 6-24 hr and thrombectomy beyond 24 hr (RR = 1.01, 95% CI [0.94, 1.07], P = 0.84) with moderate heterogeneity in the combined data (P = 0.11, I2 = 46%). (Figure 6c)

## Discussion

Our study demonstrated a possible association of EVT with better clinical outcomes in acute ischemic stroke patients presenting beyond 24 hours. This was evidenced by achieving successful recanalization in more than 80% of cases and functional independence in more than one third of the patients. Moreover, the comparison with medical treatment showed favorable outcomes in the direction of EVT and the results did not differ significantly with the regular extended window results (6-24h). These results are iconoclastic enhancing a new paradigm in which a contemporary restriction to specific time window to treat patients rather than their own clinical and imaging characteristics seems to be anecdotal. Our results are consistent with the results of four recent comparative studies that showed an association of EVT with good clinical outcomes. ^16, 20, 22, 23^ Additionally, a single arm meta-analysis including 7 studies showed encouraging results.^24^ However, our current meta-analysis included a larger number of studies and had two pairwise comparisons (EVT vs. medical management and EVT beyond 24 h vs.

EVT within 6-24 h). Hence, the results are more robust and confirmatory of the possible role of EVT beyond 24 h.

None of the studies included in our meta-analysis were RCTs which make our results prone to selection bias of cases with favorable outcome based on advanced imaging modalities. In the extended window, studies argued that selection criteria of DAWN and DFFUSE 3 should not necessarily determine the only patients can benefit from EVT.^25^ In that context, an analysis of 102 patients not consistent with DAWN and DFFUSE 3 criteria showed that EVT still has higher odds of favorable outcomes and lower odds of all-cause mortality within 90 with no significant differences in sICH rates.^5^

Real exact onset times are not clearly defined in many of the patients presenting in this very late window. For instance, in Sarraj et al^16^, around four-fifths of all included cases were of an unwitnessed onset which was also noted in extended window trials.^2, 3^ Likewise, it does not seem bizarre that some patients outside the current extended window could benefit from EVT giving the possible collateral role to maintain the benefit for those patients as slow progressors.^26^ The natural history of patients kept on medical management (only 11.3 % achieving functional independence at 90 days vs. 37% in the EVT group) could be possibly explained by failure of collateral with time in case of non-recanalization by EVT. In Sarraj et al, patients with a perfusion imaging mismatch had better clinical outcomes with EVT and significantly higher odds of functional independence (35% vs 17%; aOR, 4.17; 95% CI, 1.15-15.17; PL=0L.03). ^16^ This is also in a similar line with AURORA findings.^4^

In an early published case series including 21 patients, the criteria sounded to focus on advanced imaging characteristics for patients’ selection. Specifically, they restricted EVT to infarct volume of infarct core volume <31mL in case of National Institutes of Health Stroke Scale (NIHSS) score ≥10 or <51mLin case of NIHSS score ≥20 in age less than 80 while this was only restricted to infarct core volume <21mL with NIHSS in older cases. ^6-8^ These criteria were not consistent across included studies although most of them considered advanced imaging in patients’ selection. However, some studies did not restrict their inclusion of cases to perfusion imaging in their selection and reported positive results in EVT treated patients which may extend the generalizability.^18^ Current evidence did not point to an effect for advanced imaging based selection on clinical outcomes for late window.^27^ Moreover, we have evidence from RCTs to support EVT in large infarcts in the established window so we would benefit from future studies looking into possible role of EVT beyond 24 h in case of large infarcts.^28^

The results of our analysis included data from the posterior circulation patients. Now, the evidence from RCTs is also supporting patients treated in early and extended window for posterior circulation.^29, 30^ However, in one of the included studies, around 41.7% presented with posterior circulation stroke in which good to excellent reperfusions was achieved in > 80% but only 16.7% of them reached favorable outcome with higher mortality rate (41.7%).^19^ These results seemed contradictory with other evidence suggesting similar outcomes of patients undergoing EVT beyond 24 hours and those receiving EVT within 6–24Lhours.^8^ We encourage future studies to address this issue with larger sample size. In addition, the rate of functional independence in our study is lower numerically than patients treated in earlier time in our pooled analysis. It is consistent with the evidence suggesting a decay of favorable outcomes over time in patients treated with EVT.^31^ We also urge that the argument should be directed more towards the benefit of intervention compared with leaving the patient without EVT and to define the edge between futility and clinical effectiveness.

To our knowledge, this is the first meta-analysis assessing EVT beyond 24 hours to have a comparison against medical group and extended window groups (6-24 h). However, it was limited by the small number of studies included in these analyses. In addition, our limitations extended to include a relatively small number of included studies that were all observational with relatively small total number of patients. Some outcomes had considerable heterogeneity given the diverse set of pooled patients included from different studies. Lack of patient level data could have also limited potential subgroup analyses and confounding control. We recommend future studies to overcome those limitations.

In this meta-analysis, EVT is associated with better clinical outcomes than medical management beyond 24 hours. Prospective comparative RCTs are needed to confirm the best eligible patients for EVT in this newly proposed window extension.

## Data Availability

All data produced in the present work are contained in the manuscript

